# Assessing the usability of the new automated medication dispensation device for patients and the adherence dashboard for real-time medication monitoring for healthcare providers: a study protocol

**DOI:** 10.1101/2023.12.16.23300074

**Authors:** Tejal Patel, Christoph Laeer, Hamed Darabi, Maxime Lachance, Michelle Anawati, Marie-Hélène Chomienne

## Abstract

**Background:** Non-adherence to prescribed medication regimens can lead to suboptimal control of chronic health conditions and increased hospitalizations. Older adults may find it particularly challenging to self-manage medications due to physical and cognitive limitations resulting in medication non-adherence. While automated medication dispensing technologies may offer a solution for medication self-management among older adults, these technologies must demonstrate usability before effectiveness can be investigated and products made available for widespread use.

**Objectives:** This study will aim to measure usability, workload, and unassisted task completion rates of an automated medication dispenser and medication adherence dashboard on the Medipense portal with older adults and their clinicians, respectively.

**Methods:** This study is designed as convergent parallel mixed methods observational study with older adults and their clinicians. Usability will be examined with the use the System Usability Scale while NASA-TLX will be utilized to assess the workload of both the device and the adherence monitoring platform. Cognitive walkthrough will be utilized prior to usability testing to identify series of steps required to use the automated dispenser and adherence dashboard. The study will assess the unassisted task completion rates to successfully operate the device. Semi-structured interviews with both types of participants will provide qualitative data with which to comprehensively gauge user experience with the automated dispenser.

**Results:** The results of this study will allow us to examine usability of both the automated medication dispensing system and the adherence monitoring dashboard from older adult and heath care provider perspectives. The results of this study will highlight and address the challenges with usability that older adults and healthcare providers may face with this device and dashboard.

**Conclusions:** The results of this study will be used to optimize the usability of both the automated medication dispenser and the adherence dashboard.

## Introduction

Worldwide, the population is aging and by 2050, there will be 2.1 billion individuals older than 60 years of age, accounting for 1 out of every 5 people.[1] The increased life expectancy is resulting in an increased prevalence of chronic disease comorbidities, functional impairment and concomitant multiple medications use.[2–6] The use of multiple medications introduces new challenges such as adverse effects, drug interactions, drug-induced diseases, complex drug dosing regimens, with resulting negative impact on medication non-adherence and an increase in medication errors.[6–9] Adherence is defined as “the degree to which the person’s behavior corresponds with the agreed recommendations of a healthcare provider.”[9] Medication non-adherence is of particular importance in management of chronic conditions. Non-adherence leads to less than optimal control of chronic health conditions and significant additional costs for the Canadian healthcare system estimated at $4 billion CAD per year.[10–14] There are numerous reasons why patients do not adhere to a medication regimen. Of the five different types of factors identified,[15,16] patient and therapy related factors are of particular importance among older adults. For example, increasing use of multiple medications to treat an increasing number of multi-morbidities results in complex therapeutic regimens which directly impact medication adherence negatively.[8,16,17] Older adults, especially those who may have mild cognitive impairment, may become more forgetful which decreases medication adherence.[17] Outside of forgetfulness and short-term memory impairment, a decline in executive function may reduce the ability of cognitively impaired individuals to organize and plan medication taking activities. [18,19] In addition, older adults may also accumulate physical limitations which impact medication taking resulting in medication non-adherence. Vision impairment increases the risk of medication errors.[20] Age-related conditions such as arthritis and Parkinson’s disease impacts the ability to open vials and punch blister packs. [21] According to the World Health Organization, the rate of medication non-adherence in developed countries nears 50%.[10]

The well recognized problem of medication non-adherence has spurred the development of numerous medication adherence technologies. A recent literature review identified 78 devices capable of providing real-time monitoring of medication intake.[22] Among these devices are vials, blister packaging, pill boxes, storage boxes and injectable and inhaler devices embedded with sensors or other technology that permits real-time tracking of medication taking through opening of vials or pill box compartments, puncturing of blisters or actuation or injection.[22] These devices offer a sizeable array of features which may impact the usability of the devices by older adults based on the physical or cognitive limitations they are facing. Usability is defined as the “extent to which a system, product or service can be used by specified users to achieve a specified goal with effectiveness, efficiency and satisfaction in a specified context of use.”[23] In their examination of key usability barriers associated with the use health technolog among older adults, Wildenbos et al. proposed a framework of four key categories: cognitive, physical, perception and motivational barriers as relevant for effective and safe use of technology.[24] Within this framework, cognitive barriers such as declining working memory, spatial cognition, attention, verbal fluency and reasoning may impact errors in the use of the technology, decrease efficiency, learnability, memorability and satisfaction.[24] Physical ability barriers such as declining speed of performance, grip strength, hand-eye coordination and flexibility of joints impact errors and efficiency of use.[24] Similarly, vision acuity, contrast detection, color vision, computer literacy, self confidence also affects learnability, efficiency, errors and satisfaction with use of technology.[24] Therefore, it is imperative that usability of medication adherence technology is assessed with older adults prior to utilizing these devices to address medication non-adherence.

Unfortunately, very few studies have examined the usability of medication adherence technology in older adults. In one study, where the usability of 21 electronic medication adherence products was investigated in older adults, caregivers and healthcare professionals, usability varied widely, with mean usability scores, as measured with System Usability Scale (SUS) per product ranging from 0 to 100.[25] However, the products tested in this study included electronic blister cards, pillboxes, and prescription vials with electronic caps with a variety of features. None of these electronic medication adherence devices automatically dispensed medications.

In one of the earliest studies examining the perceived usefulness and satisfaction of an automated medication dispensing device, 96 frail older adults receiving home care used one such device, the MD.2 dispenser, for one year after which they reported on the ease of use, reliability, acceptability, routine task performance and medication management assistance.[27] The MD.2 medication dispenser was 13” by 12” by 14” machine which held 42 medication cups and could dispense 1 to 6 cups per day. The front of the machine had a delivery ramp, an alert light, an LCD message screen and a dispensing button. The dispenser dispensed the allocated medications in a cup when dispensing button was pressed. In this study, 94% of participants found the device very easy to use and 84% indicated they would use it in the future. However, the results are not reflective of initial usability, i.e. immediately after the implementation of the device The study did not address the usability concerns that frail older adults may have faced in the first days or weeks of use. Furthermore, pre-filled cups with unit doses of patient’s medications were refilled in the MD.2 every two weeks by nurses and not the participants. This further limited a true test of usability of the device by participants by limiting the interaction with the machine to one of just dispensing. Finally, participants were receiving home care and had frequent interactions with healthcare providers who assisted with the automated dispensing device.

In their study examining the impact of cognitive impairment on the usability of an electronic medication delivery device, Ligons et al., demonstrated a significant relationship between cognitive impairment, measured with Mini Mental Status Examination (MMSE) scores and percentage of task success. For example, individuals with MMSE scores of 24 and above (no cognitive impairment) were able to successfully complete 69% of the tasks compared to only 34% of those with MMSE scores of <24 (p = 0.04).[28] The automated medication dispenser used in this study was designed to deliver medications from single dose blister cards based on a schedule programmed by pharmacies. Up to 10 different blister cards could be loaded into this device and users interacted with the device through a touch screen interface. At the time of scheduled dose, the automated dispenser would beep and display a flashing message on the touch screen to alert the user to take their medication doses. Participants were then expected to confirm their readiness to take their medication as well as retrieve the blister card from the dispensing drawer, and extract the dose from the blister. The participants were tested on several usability tasks such as loading and unloading blister cards, removing pills from the blister cards, manual drop, and viewing inventory. Usability was measured with SUS and observed by researchers as they interacted with the dispenser. Only three of the 19 participants scored SUS at 80 or higher. Task success rates ranged from 10.5% for manual drop to 57.9% for viewing inventory.

In a more recent study, usability, usefulness, satisfaction and impact on caregiver burden of an automated medication dispenser was tested with 58 older adults and 11 caregivers.[28] Usability was measured with SUS and Usefulness, Satisfaction and Ease of Use questionnaire. In this study, the mean SUS scores was 85.74 (SD 12.7, range 47.5 – 100). More than 75% of participants agreed with the statement that the product was easy to use. The automated medication dispensing system, spencer, is a rectangular shaped device with a touch screen, a narrow opening in the front that allows the dispensation of single multidose medication pouch and opening at the top for loading of refill boxes. It dispenses multidose pouches of the participants’ medications at scheduled times. The multidose pouches are packaged in strips by pharmacies and supplied in boxes that are loaded into the dispenser. Pouches can be dispensed with the use of a touch screen interface. Although most participants found the automated dispenser easy to use, usability was only measured at the end of the 6 months intervention. Similar to the study conducted with the MD.2 automated dispenser, this study did not capture the usability of the device at the beginning of the intervention. Furthermore, the study did not observe the participants who successfully carried out the tasks and which required more assistance.

While all the automated medication dispensers appear to have a touch screen or LCD interface and all appear to dispence medications that are prepackaged into a multidose container, the usability of the different automated dispensers varies. Some digital interfaces may be challenging for some older adults, while others may not be able to complete the loading of refills. Therefore, it is necessary to observe how older adults interact with the automated dispenser at their initial interaction as well as after a period of time of regular use. Assessment of usability and task success at the initial interaction is necessary to identify whether the usability challenges encountered can be addressed before long-term use is implemented. Assessment of long-term usability is necessary to measure learnability of using the device appropriately to ensure safe medication dispensing.

Therefore, we aim to study the overall and task specific usability of a new automated medication dispenser with older adults at the start of the study intervention and after 6 weeks of use. We will also examine the usability of a connected real-time medication intake monitoring dashboard with the participants’ health care providers. The dashboard enables clinicians to view adherence metrics of medications their patients are prescribed while they use the automated medication dispenser. As with measuring the usability of automated dispensers, usability of accessing and viewing adherence data is also just as important to measure. If there are usability concerns with accessibility of the dashboard, or with interpretability of adherence metrics by clinicians, useful information may not be utilized in clinic to adherence medication non-adherence.

## Ethical Approval

This research project has undergone ethical review and approval by the Hôpital Montfort Research ethics board (HM-REB) ID 19-20-08-020, Ottawa, Ontario, Canada. Prior to enrolling in the study, participants will be informed of the study details in writing, and will provide written, informed consent for limited data abstraction from their medical records.

## Methods

### Study Design

This study is designed as a prospective, parallel mixed-methods convergent study. We will utilize both quantitative measures, such as the System Usability Scale [29] and NASA Load Index (NASA-TLX) [30] as well as semi-structured interviews with participants to realize the potential barriers to appropriate use of the automated medication dispenser as well as the medication adherence tracking platform. Prior to testing the usability of the automated medication dispenser and the adherence tracking platform, we will collaboratively establish steps for an appropriate use through the use of cognitive walkthroughs with the developers of the automated medication dispenser as well as the adherence dashboard, clinicians and researchers.[31] An overview of the study process can be found in Fig 1.

**Figure 1:**
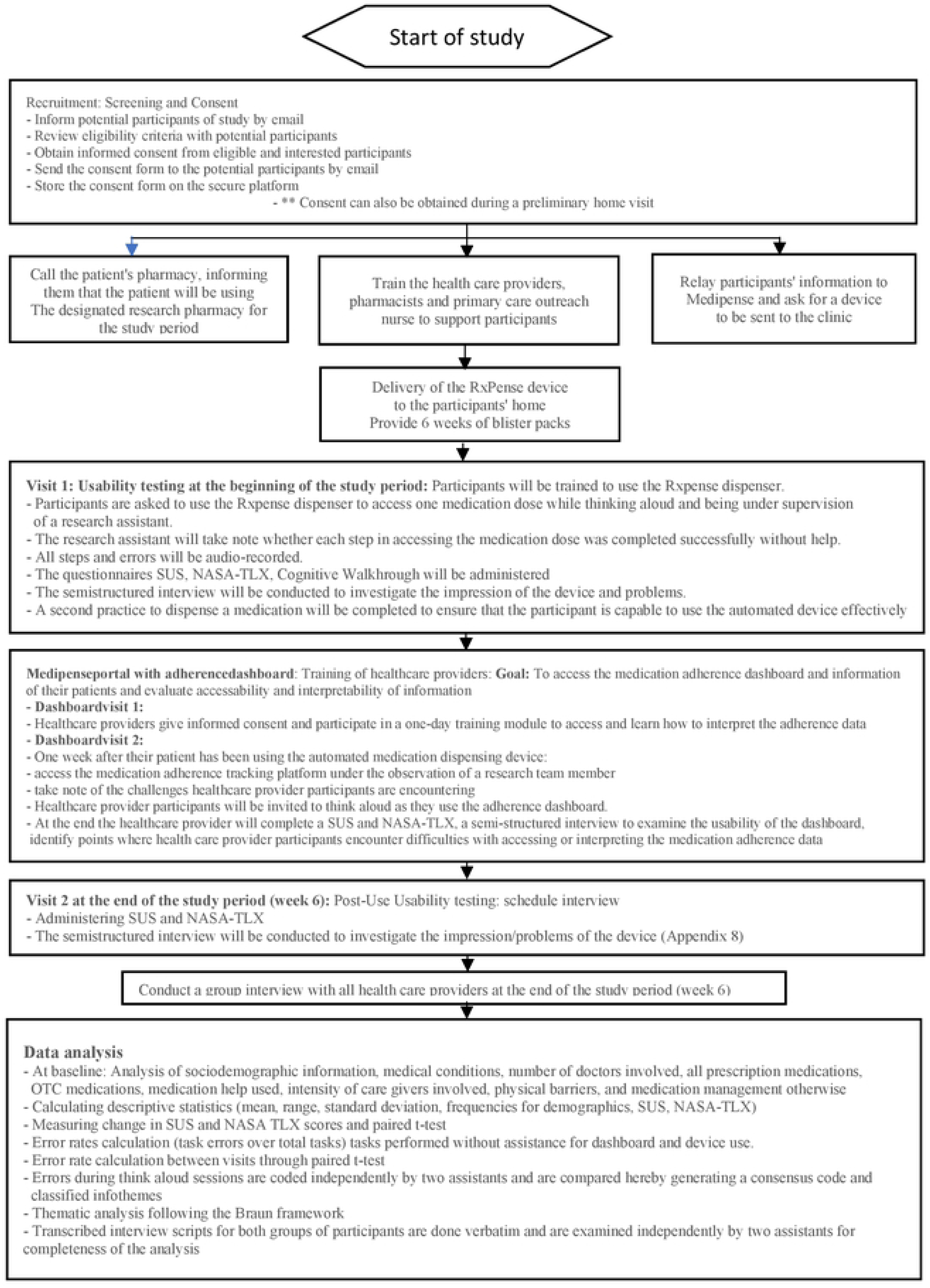
Study process

### Study Participants

#### Inclusion criteria

Older adult participants: Older adults, aged 65 years and older, will be recruited from patients presenting to one family health team in Ontario, Canada. Additional inclusion criteria include the use of three or more prescriptions and/or over-the-counter medications as well as having at least two chronic conditions.

#### Healthcare provider participants

Clinicians, including family physicians, pharmacists and nurse practitioners who are interested in accessing real-time medication adherence data on the adherence dashboard of their patients who have enrolled in the study will also be invited to participate.

#### Exclusion criteria

Older adults participants who are non-French-speaking will be excluded as the Family Health Team serves primarily a French population. Additionally, individuals with a diagnosis of dementia (any type) or cognitive impairement as noted in their electronic medical record will be excluded as this could impact participants’ ability to learn and accurately use the dispensing device.

#### Healthcare provider participants

Healthcare providers whose patients have not enrolled in this study will be excluded.

### Sampling technique

We will use purposive sampling techniques to recruit both types of participants in this study.

### Sample Size

A sample size of 5 is adequate to identify up to 80% of the usability problems with a device; [32] therefore, we will aim to recruit at least 5 and up to 10 older adults and their healthcare providers, including their family physician, nurse practitioner and pharmacist, to test the usability of the automated medication dispenser and adherence dashboard.

### Location

This study will be conducted at the Équipe de santé familiale communautaire de l’Est d’Ottawa which has two sites in Ottawa. Currently, 12 doctors, 4 nurse practitioners, 4 registered nurses, 1 psychologist, 1 social worker, 1 pharmacist, and 1 dietitian serve approximately 9000 patients (June 3^rd^, 2023).

#### Recruitment period

The recruitment period is planned from January 1^st^ 2024 to February 28^th^ 2024

### Automated Medication Dispenser

In this study, we will use Medipense RxPense® device (9.84” by 14.06” by 18.09”). [33] It is a dispensing device with a remote monitoring solution that ensures that the user will take the correct medications on time. Medications in this packaging will hold up to two weeks’ worth of medication supply with 56 individual containers delivering up to 8 doses a day. The device notifies users about the next dose by voice prompts, visual alarms, remote notifications to a wearable alert device, email or SMS alerts. If a user misses a dose the designated caregivers will be notified through SMS or email. It records and documents the use of all prescriptions including “as needed” prescriptions and over-the-counter medications. Medications are securely locked into the device and can only be dispensed when the user is authenticated at the right time through the use biometric, radio frequency identification or a password. Any missed doses will be kept in the RxPense ® device. The dispensing mechanism is arthritis friendly and the display supports visually impaired persons. Verification and audit trail is also provided with images during pill dispensing and after the patient is taking the pills. An audit trail is securely stored (HIPAA and PIPEDA compliant) in the RxPense® Cloud.

The RxPense device is connected to the RxPense® Portal and provides real time information and access to the user relevant information, monitoring and adherence by all defined members of the circle of care. [34] The RxPense® Portal allows tracking and reporting of medication adherence on an organziationsal and patient level. Before or after dispensing the medications, patients can be asked questions about their well-being, health or consumption habits. Data collected may be stored anonymized for further analysis. Reports are displayed to the patient on the RxPense Hub screen. It can also capture monitor and store vitals (through external sensors), in the electronic patient record.

### Outcomes

We aim to examine the overall usability with the System Usability Scale (SUS). The SUS has been used to examine the usability of medication adherence products in previous studies.[29] It is a quick and easily administered end-of-test subjective assessment of the usability of a product. It consists of 10 statements (5 positive and 5 negative) which are scored immediately after testing a product on a 5-point Likert scale. Scores range from 0 to 100, higher scores indicating more user-friendliness of the product. We will also examine the workload involved in using the automated medication dispenser and the adherence tracking platform. Human mental workload is an important concept associated with usability. It refers to cost associated with performing a cognitive task and can be used to predict operator and/or system performance.[35] Interaction between workload and usability drive objective performance of technology. Indeed, a previous study demonstrated that higher workload with setting up and using a medication adherence product was associated with declining SUS scores.[25] In this study, NASA-TLX will be used to measure workload.[30] The NASA-TLX consists of six subscales: mental, physical and temporal demands, frustration, effort and performance. Participants will be asked to rate each of the above-mentioned variables on a 20-point scale that measures from high to low (scored from 0-100). Finally, before we initiate the usabilty studies, we will conduct cognitive walkthrough with the automated medication dispenser and the adherence tracking platform collaboratively with the developers of the dispenser and the research team. Cognitive walkthrough methodology examines the level of difficulty in completing particular tasks within a system and will enable the researchers to identify the pain points of using both the automated dispenser and adherence dashboard by older adults and clinicians, respectively. [31] Key tasks for using both the automated dispenser and the adherence dashboard will be determined a priori by investigators. These include key tasks that are completed frequently or critical to complete and those that exhibit core capabilities of the system. Participants are then invited to “use” the automated medication dispenser or the adherence dashboard while thinking aloud and under observation by investigators; points at which the completion of key tasks fail will be noted for further development and iteration on a cognitive walkthrough checklist. Think aloud refers to participants verbalizing their thoughts as they complete the tasks required to use the automated medication dispenser or the adherence dashboard.[36] Think aloud enables researchers to gain insight into what the participant is thinking and reflect on why errors occur. Unassisted completion rates of all tasks (number of steps completed accurately without assistance/total number of steps) and key errors (and reasons for these errors) will be reported.[37]

### Intervention

Patients who agree to participate will test the device at home. All participants will be trained by the developers of the automated dispensing system. A nurse will provide home support to participants. The duration of the usability study will be 6 weeks per older adult participant. We plan 2 visits for all participants. During the first visit at the beginning of the study period, all participants will provide informed consent and will be trained to use the automated medication dispenser. Following the training, participants will be asked to use the automated dispenser to access one dose of their medication regimen while thinking aloud and being under observation by a research team member. As a part of the cognitive walkthrough, participants will be observed by a member of the research team during the use of the dispenser. A research member will note whether each step in accessing the medication dose is completed successfully without assistance. These sessions will be audio recorded. If the participant encounters any problems with successfully completing the steps, these will be noted in detail by the research assistant on the Cognitive Walkthrough data collection sheet. Once this medication has been dispensed, they will be asked to complete SUS and NASA-TLX. Following the completion of both tools, participants will participate in a one-on-one semi-structured interview designed to investigate their impression of the device, the particular problems they encountered with the dispenser, and discuss any assistance they required to successfully complete the task of dispensing the medication. Once the interview is completed, researchers will complete a second round of training with the participant to ensure they are able to use the automated dispenser effectively for the remainder of the study duration. At the end of the 6-week duration of the study, we will complete a second visit (Visit 2) with the older adult participants. During this visit, participants will complete the SUS and NASA-TLX again. Another semistructured interview will be conducted to investigate the participants’ experience with using the device over the 6 week period.

Healthcare providers whose patients have agreed to participate in the study and who are interested in accessing the medication adherence dashboard will be invited to participate in a one-day training module where they will be trained on the steps required to access the adherence data for their patient as well as to intepret the information available. During this training session, they will also be asked to provide informed consent. Once they are trained, they will be provided with access to their patient’s medication adherence information. One week after their patients have been using the automated medication dispensing device, they will be asked to access the medication adherence tracking platform while under the observation of a research team member, who will note the challenges the healthcare provider encounters while accessing and using the platform. Similar to the older aldult participants, healthcare provider participants will be invited to think aloud as they use the adherence dashboard. At the end of this session, the healthcare provider will complete a SUS and NASA-TLX and participate in a semi-structured interview designed to further examine the usability of the dashboard, identify points where clinicians encounter difficulties with accessing or interpreting the medication adherence data. At the end of the six week study period a semi structured group interview with all health care providers will be conducted to gather further information about the device experience.

### Data collection

In addition to SUS and NASA-TLX, we will also collect socio-demographics (age, self-identified gender, sex at birth, medical conditions, name and dosing regimen of prescribed and “as needed” prescription and over-the-counter medications, number of doctors and other healthcare providers involved in the patient’s care, any physical or sensory barriers (for example, physical strength of upper extremities, pain, numbness, tremor in upper extremities, vision or hearing impairment) to medication management. We will ask participants to identify if they use any medication taking aids as well as how long they have used these aids. We will capture the type and intensity of caregiver support that older adult participant have access to at home. Demographic data (age, self-identified gender, sex at birth, years of practice, discipline, patient roster size) for healthcare providers participating in this study will also be collected.

### Analysis

Descriptive statistics (mean, range, standard deviations and/or frequencies) will be reported for quantitative measures (demographic data, SUS and NASA-TLX). Change in SUS and NASA-TLX scores between the first and second visits among participants using the automated medication dispenser for 6 weeks will be examined for significance with a paired t-test. Statistical analysis will be conducted with the use of R. [30]

Error rates will be calculated by dividing the number of tasks errors made by each participant while using the automated medication dispenser or the adherence dashboard divided by the total number of steps required to use the dispenser or dashboard. Unassisted task completion rates will be measured by dividing the total number of tasks completed by each participant without assistance while using the dispenser or dashboard. Error rates, unassisted completion rates between the two visits will be compared with a paired t-test. The errors identified during the think aloud sessions will be coded qualitatively and classified into themes for each task.

Think aloud sessions and semi-structured interviews for both groups of participants will be audio-recorded, transcribed verbatim and examined for completeness and accuracy by two independent research team members prior to the initiation of data analysis The framework by Braun et al [38] will be used to perform thematic analysis for the semi-structured interviews. Two transcripts for each of the participant types will be coded independently by two research team members to identify preliminary codes. These codes will be compared between the two transcribers to resolve any discrepancies, develop a consensus on codes and their definitions/meaning and finalize a coding manual. This coding manual will be used as a reference for coding the remaining transcripts by one research team member for each type of participant independently. Codes that are generated will be classified into themes.

## Impact

Completing the pilot usability study for automated medication dispensing device will permit a determination of whether older adults are able to use the device for its intended purpose at home while examining the usability of the real-time medication monitoring and adherence portal is importance to identify whether healthcare clinicians are able to easily access the adherence data for the management of their patients. This study will guide the implementation of the automated medication dispensing device in primary care by ensuring that older adults are able to use the device appropriately at home as well as ensure that clinicians use the adherence data available to manage medication non-adherence. The study results will also allow the developers to optimize the functionality of the automated medication dispensing device and the adherence monitoring platform to meet the needs of both older adult and clinician end-users.

## Discussion

Our patient population will be using three or more chronic medications and we will test the device for a 6-week period at home. We expect that the results will reflect the feasibility of the usability, acceptance, and workload of the device from the patient’s perspective. In addition, it will assess three things: the usability of the RxPense platform for health care providers, patients and, potentially, care givers; real time adherence monitoring from a health care provider perspective; and integration of this system into the primary care sector. Real time monitoring of adherence will allow caregivers a better understanding of the factors related to non adherence as well as the opportunity for early interventions. Objective, in time adherence information on the patient or aggregate level will help clinicians to unmask and understand the dimensions of non-adherence. It will create a shame- and blame-free environment to ask questions as objective information are discussed. The relationship between clinician communication and adherence has been studied since the 1960s [39]. A Meta-analysis by Zolnierek and colleagues in 2009 point out that physician communication is significantly positively correlated with patient adherence and that there is a 19% higher risk of nonadherence among patients whose physician communicates poorly than among patients whose physician communicates well. [40] Enhanced communication and interaction between the prescriber and the patient will allow to unmask various patient behaviors that can be addressed directly and help to better understand real life at home barriers to adherence. Studies show that patients make changes to their prescription regimen (e.g., adjusting doses, or times) whilst withholding the information from their health provider [41,42,43] Improved knowledge of the changes may allow more conversations with the health provider towards improved shared decision-making, or increase needed patient education and information on their medications or prescription regimen towards better self-management. The information collected will also have the potential to inform the health provider on side effects the patient may be encountering. Thus improving medication adherence. On the aggregated data level, a tailored approach for adherence specific communication processes and routinely asking questions can be developed and can involve patients. [43]

### Strengths of the Study

The study answers a need to assist patients living with comorbidities who take multiple medications. Adults rarely report the problems they have with medication management. Our study assesses the usability of an innovative device and digital platform from a patient and provider perspective. The patients are recruited from a team-based primary care interdisciplinary environment serving a middle income active population (the majority of the population is between 15 and 65 years of age) in suburban Ottawa, Ontario. The study uses standardized, established usability assessment tools as well as targeted questions and semi-structured interviews to understand the system challenges. It assesses adherence in real time directly in the home of the patient. An advantage of a study of this system is that it provides guidance to future patients who must navigate complex multidrug regimens by eliminating the need for patient decision-making concerning what medication to take, how much, and at what time thereby improving adherence. Further on, the dispenser mechanism will prevent patient medication overadherence and administration of medication at incorrect time intervals.

The system possesses the ability to wirelessly transmit patient medication adherence data, providing opportunities to assess and monitor patient medication adherence in real time. The information can be interpreted by the patient, care giver and health care provider. In addition, health care providers can interpret information of adherence challenges and attempt to identify specific population characteristics to include the learning in adherence improvement measures.

### Limitations of the Study

Given the novel aspect of the study, the project might highten the participant’s anxiety. Participants are asked to test a product they have not been exposed to before for essential health matters, and this could negatively impact the results. In addition, because the opening of the device dispensing drawer is used as a proxy measure for adherence, patient actions such as failing to ingest removed medications can lead to inaccurate estimates of patient medication adherence and raise concerns toward their medication adherence monitoring accuracy because of potential patient behaviors.

## Data Availability

Deidentified research data will be made publicly available when the study is completed and published.

## Credit Authorship Contribution Statement

Tejal Patel: Conceptualization, methodology, writing – original draft, writing – review & editing Christoph Laeer: Conceptualization, methodology, writing – original draft, writing – review & editing, project administration, supervision

Marie-Hélène Chomienne: Conceptualization, methodology, writing – original draft, writing – review & editing

Hamed Darabi: Project administration

Maxime Lachance: Project administration

Michelle Anawati: Funding aquisition, conceptualization, writing – review and editing

## Acknowledgements

We are grateful to the ESFCEO administration, the providers and board of directors for their support. We would like to thank Dr. Sadaf Faisal and Ms. Jessica Ivo for assisting with study methodology. We would like to thank Terry Fagon and the whole Medipense team for their support, granting access to the RxPense Portal and the device in planning this study.

## Supporting information

S1 Fig Study process

